# External Validations of Cardiovascular Clinical Prediction Models: A Large-scale Review of the Literature

**DOI:** 10.1101/2021.01.19.21250110

**Authors:** Benjamin S. Wessler, Jason Nelson, Jinny G. Park, Hannah McGinnes, Gaurav Gulati, Riley Brazil, Ben Van Calster, D. van Klaveren, Esmee Venema, Ewout Steyerberg, Jessica K. Paulus, David M. Kent

**Affiliations:** Predictive Analytics and Comparative Effectiveness (PACE), Tufts Medical Center, United States of America; Division of Cardiology, Tufts Medical Center, Boston, MA; KU Leuven, Department of Development and Regeneration, Leuven, Belgium; Department of Biomedical Data Sciences, Leiden University Medical Centre, Leiden, Netherlands; Department of Public Health, Erasmus MC University Medical Center, Rotterdam, the Netherlands; Department of Neurology, Erasmus MC University Medical Center, Rotterdam, the Netherlands; Department of Biomedical Data Sciences, Leiden University Medical Center, Leiden, Netherlands

## Abstract

**Background:** There are many clinical prediction models (CPMs) available to inform treatment decisions for patients with cardiovascular disease. However, the extent to which they have been externally tested and how well they generally perform has not been broadly evaluated.

**Methods:** A SCOPUS citation search was run on March 22, 2017 to identify external validations of cardiovascular CPMs in the Tufts PACE CPM Registry. We assessed the extent of external validation, performance heterogeneity across databases, and explored factors associated with model performance, including a global assessment of the clinical relatedness between the derivation and validation data.

**Results:** 2030 external validations of 1382 CPMs were identified. 807 (58%) of the CPMs in the Registry have never been externally validated. On average there were 1.5 validations per CPM (range 0-94). The median external validation AUC was 0.73 (25^th^ −75^th^ percentile [IQR] 0.66, 0.79), representing a median percent decrease in discrimination of −11.1% (IQR −32.4%, +2.7%) compared to performance on derivation data. 81% (n = 1333) of validations reporting AUC showed discrimination below that reported in the derivation dataset. 53% (n = 983) of the validations report some measure of CPM calibration. For CPMs evaluated more than once, there was typically a large range of performance. Of 1702 validations classified by relatedness, the percent change in discrimination was −3.7% (IQR −13.2, 3.1) for ‘closely related’ validations (n=123), −9.0 (IQR −27.6, 3.9) for ‘related validations’ (n=862) and −17.2% (IQR −42.3, 0) for ‘distantly related’ validations (n=717) (p<0.001).

**Conclusion:** Many published cardiovascular CPMs have never been externally validated and for those that have, apparent performance during development is often overly optimistic. A single external validation appears insufficient to broadly understand the performance heterogeneity across different settings.

## Introduction

Clinical prediction models (CPMs) are widely available to inform decisions in cardiovascular medicine. Our own database, the Tufts Predictive Analytics and Comparative Effectiveness (PACE) CPM Registry,^1^ demonstrates continued growth of prediction models for patients with cardiovascular disease (CVD) despite apparent substantial redundancy. The growth in the literature reflects the increasing ease with which these models can be developed, given the wide availability of both data and statistical software. Despite the publication of methodologic^2^ and reporting guidelines^3^ and a large set of potential performance metrics^4^, much remains unknown about the broad performance of these models, including the extent to which they have been validated, how well they validate, and how performance varies from one setting to another.

While there are various ways to assess the performance of a statistical model^4^, clinically beneficial CPMs will yield accurate predictions on new cohorts (external validation)^5^ and improve decision making and subsequent clinical outcomes. Despite the increasing number of CPMs in the literature, how models perform generally during external validations and the determinants of that performance is largely unknown. Current reporting recommendations reinforce the need for external validation^3^ though recent analyses suggest that most CPMs either have not been externally validated^6^ or have only been validated on a single external cohort.^7^ CPM discriminatory performance cannot not be assumed to be stable (i.e. equivalent to model performance at derivation) when tested in new settings.^8^ Model calibration has been largely neglected and unless it is known to be excellent, CPMs may lead to harm if they are used to inform decisions at certain risk thresholds^9,10^.

Here, we perform a field synopsis of external validation studies of cardiovascular CPMs reported in a prior systematic review.^1^ We aimed to describe the extent of external validation, variation in performance of models across databases, and to explore factors that are associated with worse model performance.

## Methods

### Cardiovascular CPMs

The cardiovascular CPMs that form the basis of this review are found within the Tufts PACE CPM Registry. This Registry is available at www.pacecpmregistry.org and represents a field synopsis of prediction models for patients at risk for and with known cardiovascular disease. The search strategy and inclusion criteria have been previously reported.^1^ Briefly, for inclusion in the Registry, an article must present the development of a cardiovascular CPM, contain a model predicting a binary clinical outcome, and the model must be presented in a way that allows prediction of outcome risk for a future patient. The search strategy for CPM identification was previously reported^1^ and is presented in the **Supplement Fiigure 1**. This analysis looked at cardiovascular CPMs published from 1990 through March 2015.

### External Validation Search

A SCOPUS citation search of these cardiovascular CPMs was conducted on March 22, 2017. Citations were reviewed by two members of the study team to identify external validations of CPMs in the Registry. Discrepancies were reviewed by a third member of the research team. Consistent with prior work^6^, external validations were defined as any report that claimed to study the CPM for the same outcome as originally reported, but in a non-overlapping population.

### Data Extraction

Information about each CPM/validation pair was extracted, including sample size, continent of study, number of events, and reporting of measures of discrimination and calibration. CPM validation performance focused on discrimination (AUC) change compared to the AUC seen in the derivation population. We also document whether validations include any assessment of CPM calibration. There are many methods to assess model calibration and only recent consensus on best practices.^4,11^ Given this lack of consistency and interpretability in the literature, we report whether or not this dimension of performance was assessed during external validation. Calibration assessment included any comparison of observed versus expected outcomes. Examples include a Hosmer-Lemeshow statistic or calibration plot. For this study we also included measures of calibration-in-the-large, where overall observed event rates are compared to predicted rates.

### CPM performance

Consistent with prior work^12^, percent changes in CPM discrimination from derivation to validation are described on a scale of 0% (no change in discrimination) to −100% (complete loss of discrimination) because it more intuitively reflects the true changes in discriminatory power.^13^ Positive changes represent improvements in discrimination. The percent change in discrimination is calculated using the following equation [(Validation AUC − 0.5) – (Derivation AUC - 0.5) / (Derivation AUC - 0.5) ∗ 100].

### Population Relatedness

To explore potential explanations for decreased performance on validation data sets, we assessed the similarity between the derivation and validation populations by creating detailed relatedness rubrics for the 10 index conditions with the greatest number of CPMs **(Supplement Taable 1)**. These rubrics were created by investigators with expertise in these clinical areas. Relatedness was assessed for each CPM/validation pair to divide validation databases into 3 categories —“closely related,” ”related” and “distantly related.” A fourth category “no match” was assigned to validations that were excluded from the analysis because they were not clinically appropriate matches (e.g., CPM validated on population with non-overlapping index condition or outcome). Generally, the relatedness rubrics were based on 5 domains: (1) recruitment setting (e.g., outpatient vs emergency room vs inpatient), (2) major inclusion/ exclusion criteria, (3) intervention type (e.g., percutaneous coronary intervention versus thrombolysis for acute myocardial infarction), (4) therapeutic era, (5) follow-up time. Two clinicians reviewed these domains for each CPM/validation match and assigned a relatedness category. Non-random split-sample validations were labeled as “closely related” validations. Discrepancies were reviewed by the study team to arrive at a consensus.

### Factors associated with CPM external validation

We identified a set of study level factors to evaluate associations with whether or not a CPM was externally validated. These factors were identified based on observed methodologic and reporting patterns as well as prior literature.^8^ These factors included: Index clinical condition, internal validation performed, year of publication (divided here before 2004, 2004-2009, 2009-2012, after 2012), continent of origin, study design (e.g., clinical trial vs. medical record), sample size, number of events, number of predictors, prediction time horizon (< 30 days, 30-265 days, >365 days), regression method (e.g., logistic regression vs Cox regression), and reporting of discrimination or calibration. We analyzed unadjusted associations and used multivariable logistic regression to assess whether these variables were associated with CPM external validation.

### Factors associated with poor performance

A set of study level factors defined *a priori* were evaluated for association with worse CPM performance (discrimination) during validation. These factors included: population relatedness (here, dichotomized as distantly related versus other), presence of overlapping authors, same or different paper, CPM modeling method, CPM data source, validation data source, outcome rate difference between derivation and validation data (defined as > versus ≤ 40%), CPM events per included variable (EPV). We used generalized estimating equations (GEE)^14,15^ with robust covariance estimator to assess the multivariable association with the observed change in discrimination, taking into account the correlation between validations of the same CPM. Multiple imputation of 20 imputed data sets was used to account for missingness. These analyses estimated the absolute difference in the estimated percent change in the c-statistic from derivation to validation populations, as calculated above. All statistical analyses were performed using SAS Enterprise Guide version 8.2 (SAS Institute Inc., Cary, NC, USA).

## Results

### Overview of Validations

The Registry includes 1382 CPMs for CVD and the citation search of these CPMs identified 54,086 citations that were screened (**Figure 1**). These citations identified 14,615 abstracts that were screened to identify 6039 full text articles. A total of 2030 external validations were extracted from 413 papers. Only 575 (42%) of the CPMs in the Registry have ever been validated. On average there were 1.5 validations per de novo CPM, with a very skewed distribution. The Logistic EuroSCORE^16^ has been externally validated 94 times. For this analysis, we included 1846 validations of 556 CPMs after exclusion of 19 decision trees and 156 validations performed on ‘unrelated’ (i.e. populations with different index conditions or non-overlapping outcomes) samples. The median external validation sample size was 861 (25^th^-75^th^ percentile [IQR] 326, 3306) and the median number of outcome events was 68 (IQR 29, 192) (**Table 2**).

**Table 1.**
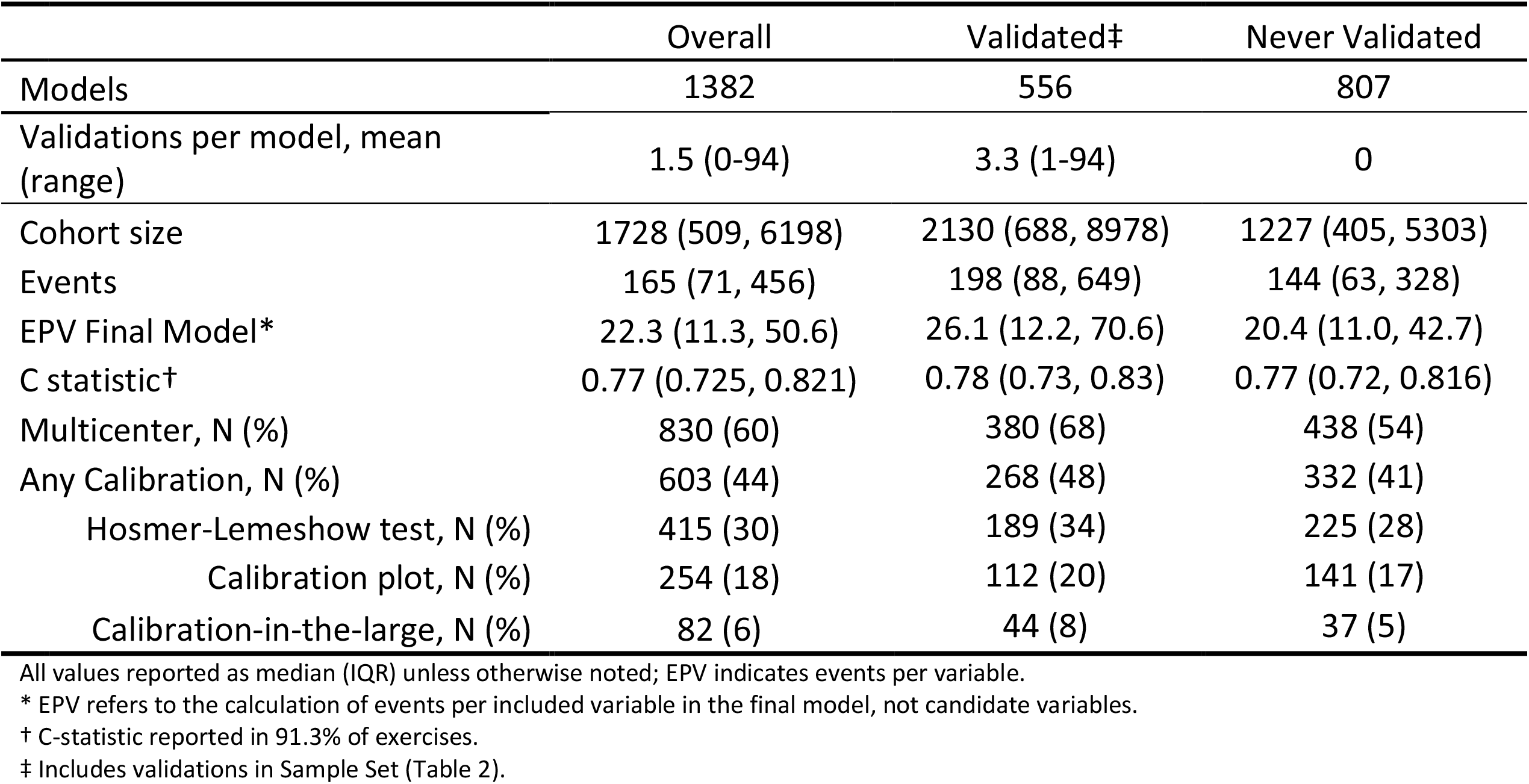
De Novo Models Summary. Characteristics of unique CPMs in PACE CPM Registry in aggregate for all CPMs, CPMs that have ever been validated, and CPMs never validated.

|  | Overall | Validated‡ | Never Validated |
| --- | --- | --- | --- |
| Models | 1382 | 556 | 807 |
| Validations per model, mean (range) | 1.5 (0-94) | 3.3 (1-94) | 0 |
| Cohort size | 1728 (509, 6198) | 2130 (688, 8978) | 1227 (405, 5303) |
| Events | 165 (71, 456) | 198 (88, 649) | 144 (63, 328) |
| EPV Final Model* | 22.3 (11.3, 50.6) | 26.1 (12.2, 70.6) | 20.4 (11.0, 42.7) |
| C statistic† | 0.77 (0.725, 0.821) | 0.78 (0.73, 0.83) | 0.77 (0.72, 0.816) |
| Multicenter, N (%) | 830 (60) | 380 (68) | 438 (54) |
| Any Calibration, N (%) | 603 (44) | 268 (48) | 332 (41) |
| Hosmer-Lemeshow test, N (%) | 415 (30) | 189 (34) | 225 (28) |
| Calibration plot, N (%) | 254 (18) | 112 (20) | 141 (17) |
| Calibration-in-the-large, N (%) | 82 (6) | 44 (8) | 37 (5) |

All values reported as median (IQR) unless otherwise noted; EPV indicates events per variable.
\* EPV refers to the calculation of events per included variable in the final model, not candidate variables.
† C-statistic reported in 91.3% of exercises.
‡ Includes validations in Sample Set (Table 2).

**Table 2.**
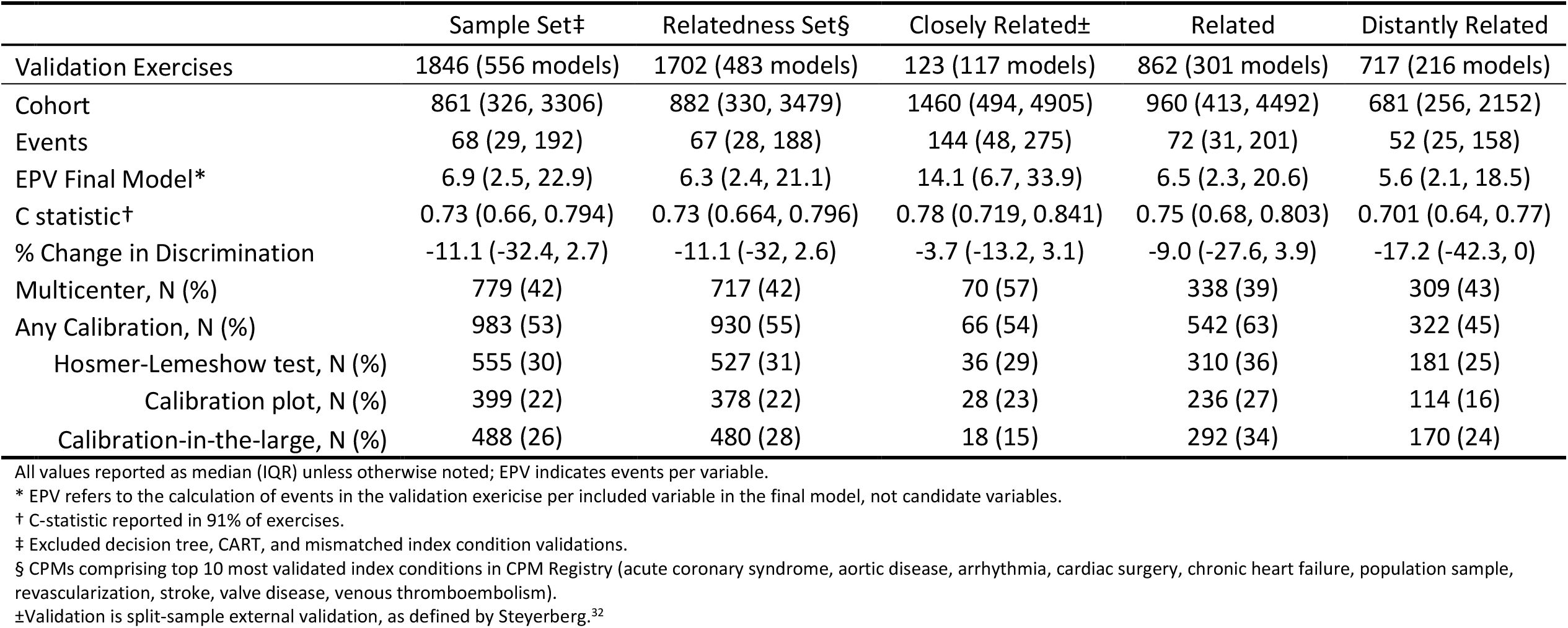
External Validations Summary. Characteristics of external validations of CPMs in PACE CPM Registry, stratified by inclusion in analysis sample, CPMs in top 10 most validated index conditions, and by relatedness category.

**Figure 1.**
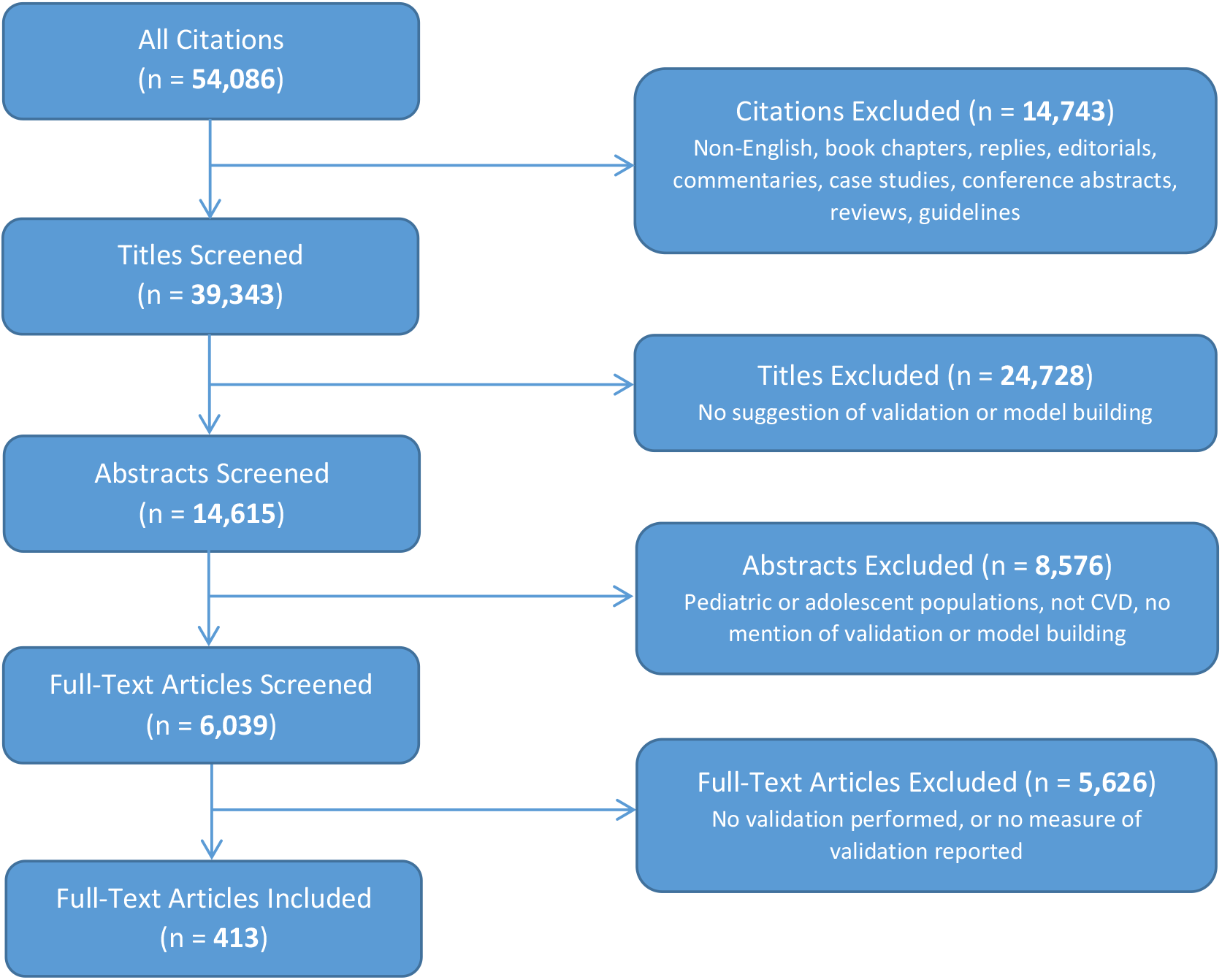
Flowchart of clinical prediction model external validation review process.

### CPM Validation Discrimination

Overall, 91.3% (n = 1685) of the external validations report area under the receiver operating characteristic curve (AUC). The median derivation AUC was 0.77 (IQR 0.73, 0.82). The median external validation AUC was 0.73 (IQR 0.66, 0.79) representing a median percent change in discrimination of - 11.1% (IQR −32.4%, +2.7%) (**Table 2**). Of the validations with decreased performance (n = 795), 25% (n = 195) had less than 10% decrement in discrimination. Two percent (n = 35) had greater than 80% drop in discrimination; 19% (n = 352) of model validations showed CPM discrimination at or above the performance reported in the derivation dataset.

### CPM Calibration

In total, 53% (n = 983) of the validations report some measure of CPM calibration. The Hosmer-Lemeshow test of goodness-of-fit was most commonly reported (30%, n = 555) followed by calibration-in-the-large (26%, 488), and calibration plots (22%, n = 399). (**Table 2**). Overall, there was no externally assessed calibration information available for 86% (n = 1182) of the CPMs in the Registry.

### Clinical Domains

The ten conditions with the most CPM validations comprised 92% (1702/1846) of the total validations included in this analysis (**Table 3**). The condition with the largest number of validations was Stroke (299 validations performed on 104 CPMs). There were a total of 286 validations of 87 CPMs for populations at risk for developing CVD (population samples) and 286 validations of 52 CPMs for Cardiac Surgery.

**Table 3:** Conditions with the Most External Validations (Top 10). Discrimination and characteristics of validations for CPMs with top 10 most validated index conditions in PACE CPM Registry.

| Index condition | Validated CPMs (% of total) | Validations | Closely Related | Related | Distantly Related | N missing | Delta C, median (Q1,Q3) |  |  |
| --- | --- | --- | --- | --- | --- | --- | --- | --- | --- |
|  |  |  |  |  |  |  | Closely Related | Related | Distantly Related |
| Stroke | 104 (48) | 299 | 5 (1.7) | 127 (42.5) | 167 (55.9) | 69 | -7.1<br>(-12.8, 2.8) | -6.9<br>(-17.7, 3.5) | -12.9<br>(-33.3, 0.4) |
| Cardiac Surgery | 52 (51) | 286 | 19 (6.6) | 216 (75.5) | 51 (17.8) | 141 | 5.9<br>(-26.9, 8.9) | -10.3<br>(-27.6, 6.9) | -17.2<br>(-43.4, 2.9) |
| Population Sample | 87 (38) | 286 | 7 (2.5) | 162 (56.6) | 117 (40.9) | 162 | -8.7<br>(-13.3, -3.6) | -15.2<br>(-38.2, -1.1) | -16.1<br>(-48.6, 1.3) |
| ACS | 57 (45) | 209 | 20 (9.6) | 85 (40.7) | 104 (49.8) | 59 | -2.3<br>(-10.3, 4.0) | -3.2<br>(-17.8, 7.8) | -11.7<br>(-39.1, 2.2) |
| Valve Disease | 37 (62) | 202 | 12 (5.9) | 71 (35.2) | 119 (58.9) | 56 | -8.9<br>(-17.9, 0) | -6.3<br>(-32.5, 2.3) | -31.8<br>(-52.3, -11.5) |
| Arrhythmia | 17 (81) | 98 | 3 (3.1) | 55 (56.1) | 40 (40.8) | 11 | -12.7<br>(-13.6, -11.8) | -17.3<br>(-50.0, 44.8) | -31.8<br>(-70.1, 46.2) |
| CHF | 47 (32) | 92 | 18 (19.6) | 47 (51.1) | 27 (29.4) | 30 | -3.7<br>(-13.5, 3.5) | -21.3<br>(-29.4, -5.5) | -17.2<br>(-28.9, -5.2) |
| Revascularization | 47 (36) | 92 | 27 (29.4) | 37 (40.2) | 28 (30.4) | 21 | -1.1<br>(-14.9, 1.6) | -1.7<br>(-16.7, 2.6) | -15.1<br>(-25.0, -5.9) |
| Aortic Disease | 31 (50) | 72 | 7 (9.7) | 32 (44.4) | 33 (45.8) | 65 | NA | -7.9<br>(-10.3, -0.9) | 39.4<br>(39.4, 39.4) |
| VTE | 27 (53) | 66 | 5 (7.6) | 30 (45.5) | 31 (47.0) | 34 | -10<br>(-17.2, -5.0) | -3.4<br>(-19.2, 28.6) | -7.4<br>(-25.0, 6.2) |
| Total | 506 (44) | 1702 | 123<br>(7.2%) | 862<br>(50.6%) | 717<br>(42.1%) | 648 | -3.7<br>(-13.3, 3.5) | -9<br>(-27.6, 3.9) | -17.2<br>(-42.4, 0) |

CPMs indicates clinical prediction models; ACS, acute coronary syndrome; CHF, chronic heart failure; NA, not applicable; VTE, venous thromboembolism. N missing refers to the number of CPMs not reporting a baseline AUC. N missing refers to either derivation AUC or validation AUC missing (delta C not available).

Only five index conditions had ≥ 50% of available CPMs externally validated [Arrhythmias (81%), Valve Disease (62%), Venous Thromboembolism (53%), Cardiac Surgery (51%), and Aortic Diseases (50%)]. There is an extreme range of CPM performance and consistent loss of discriminatory performance during external validations (**Figure 2, Table 3**). These observations were apparent for all conditions that were studied (specific condition waterfall analyses shown in **Supplemental Figure 2**).

**Figure 2.**
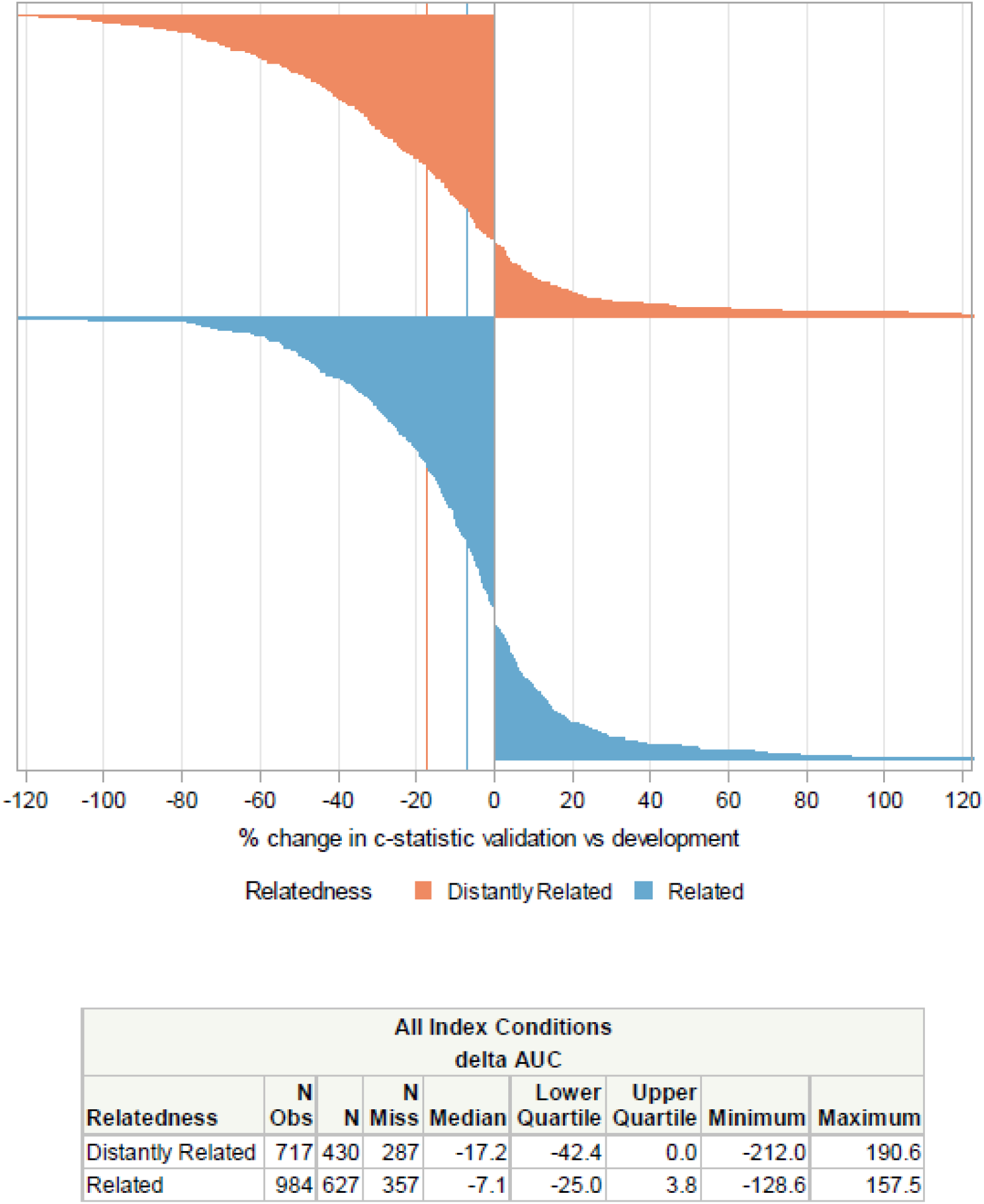
Percent Change in Validation C-statistic Performance versus Derivation C-statistic Performance. Waterfall plot depicting the percent change in the c-statistic in related [related and closely related] validations (in blue) and distantly validations (in orange). Vertical lines show that the median decrement in discrimination was more pronounced in the distantly related models than the related models.

### Relatedness

Relatedness was assigned to each of the 1702 of the CPM/validation pairs for the top 10 index conditions. Of these, 123 (7%) of the validations were performed on ‘closely related’ populations, 862 (51%) were performed on ‘related’ populations, while 717 (42%) were performed on ‘distantly related’ populations **(Table 2)**. The median AUC for ‘closely related’ validations was 0.78 (IQR 0.719, 0.841). The median AUC for ‘related population’ validations was 0.75 (IQR 0.68, 0.803). The median AUC for ‘distantly related’ validations was 0.70 (IQR 0.64, 0.77) (p <0.001). Overall, the median percent change in discrimination was −3.7% (IQR −13.2, 3.1) for ‘closely related’ validations, −9.0 (IQR −27.6, 3.9) for ‘related validations’ and −17.2% (−42.3, 0) for ‘distantly related’ validations (p<0.001).

### Range of Performance for Individual CPMs

**Table 4** shows the variation in performance across the 10 CPMs that were validated most frequently. Uniformly, there was a substantial range in performance of each CPM across datasets, from virtually useless to excellent. For example, discrimination for the Logistic EuroSCORE (validated 94 times) ranged from 0.48 to 0.90 across different databases. None of these highly cited (and validated) CPMs had consistently good discrimination across validation databases.

**Table 4.** Top 10 Most Validated CPMs. Description of top 10 most validated CPMs in PACE CPM Registry and validation performance.

| Model Name | Index Condition | Number of validations | Development AUC | Median validation AUC (IQR) | Range in validation AUC |
| --- | --- | --- | --- | --- | --- |
| Logistic EuroSCORE | Cardiac Surgery | 94 | NR | 0.75 (0.67, 0.80) | 0.48-0.90 |
| Additive EuroSCORE | Cardiac Surgery | 86 | 0.79 | 0.77 (0.72, 0.82) | 0.58-0.90 |
| EuroSCORE II | Valve Disease | 65 | 0.81 | 0.76 (0.68, 0.81) | 0.40-0.87 |
| GRACE | CAD: ACS | 53 | 0.83 | 0.80 (0.73, 0.84) | 0.60-0.95 |
| STS (valve) - Mortality | Cardiac Surgery | 51 | 0.81 | 0.70 (0.64, 0.76) | 0.45-0.85 |
| CHA<sub>2</sub>DS<sub>2</sub>-VASc | Arrhythmia | 45 | 0.61 | 0.66 (0.61, 0.69) | 0.45-0.93 |
| CHADS<sub>2</sub> | Arrhythmia | 37 | 0.82 | 0.65 (0.61, 0.68) | 0.51-0.87 |
| FRS - CHD | Population Sample | 35 | NR | 0.68 (0.63, 0.72) | 0.54-0.80 |
| ICH Score | Stroke | 27 | 0.92 | 0.85 (0.75, 0.87) | 0.69-0.94 |
| ACEF Score | Cardiac Surgery | 26 | 0.74 | 0.74 (0.68, 0.77) | 0.54-0.87 |

CPMs indicates clinical prediction models; AUC, area under curve; IQR, interquartile range; EuroSCORE, European System for Cardiac Operative Risk Evaluation; NR, not reported; CAD, coronary artery disease; ACS, acute coronary syndrome; GRACE, Global Registry of Acute Coronary Events; STS, Society of Thoracic Surgeons; CHADS-VASc, Congestive heart failure/LV dysfunction, Hypertension, Age, Diabetes mellitus, Stroke/TIA/TE, Vascular disease, Age, Sex category; CHADS, CHF, Hypertension, Age, DM, History of Stroke or TIA; FRS – CHD, Framingham Risk Score for Coronary Heart Disease; ICH, IntraCerebral Hemorrhage; ACEF, Age, Creatinine, and left ventricular Ejection Fraction.

### Predictors of External Validation

Study features that are associated with CPM external validation (yes/no) are shown in **Supplemental Table 2**. The index condition was strongly associated with subsequent external validation. Models that were internally validated and models that were published more recently were less likely to be externally validated. Sample size, number of predictors, and reporting of discrimination or calibration were positively associated with external validation. On multivariable analysis, these predictors remained associated with CPM external validation. Study design, prediction time horizon, and regression method were not associated with a model being externally validated.

### Predictors of Poor Performance

Predictors of CPM validation performance are shown in **Table 5**. On univariate analysis, population relatedness was significantly associated with CPM discrimination in validations. When CPMs were tested on “distantly related” cohorts, the AUC decrease was −15.6% (95% CI −22.0, −9.1) compared to the reference (validations done on “closely related” cohorts). When evaluated in a multivariable model, population relatedness remained significantly associated with CPM discrimination in validations (−9.8%; 95% CI −18.8, −0.8). We also observed that validations demonstrated AUCs that were 9.8% (95% CI 5.4, 14.2) higher when reported in the same manuscript (with the same authors) as the de novo CPM report compared to validations reported in different manuscripts with non-overlapping authors.

**Table 5.**
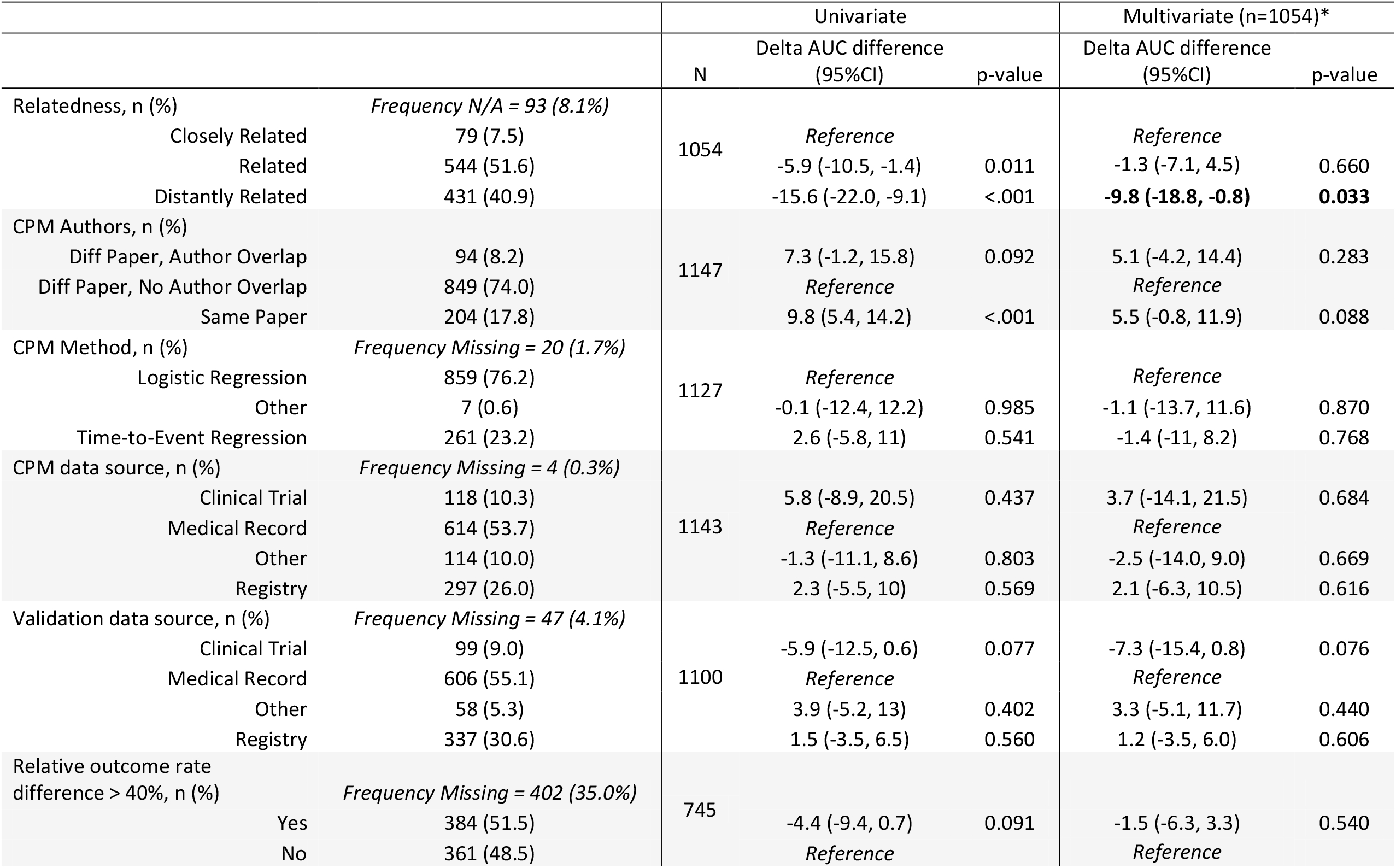
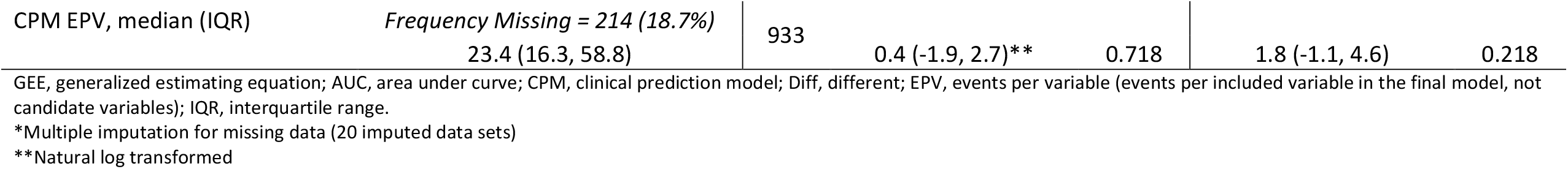
Predictors of Worse Discrimination: Variable distributions and GEE model results. Results of regression analysis to detect predictors of change in discrimination performance from derivation to validation.

## Discussion

Our Tufts PACE CPM Registry documents the tremendous proliferation and redundancy of CPMs being developed and published. The review reported here underscores that this proliferation is occurring without adequate—or even minimal—external evaluation. Approximately 60% of published CPMs have never been externally validated. Approximately half of the CPMs that have been validated have been validated only once. A small minority of models have been validated numerous times. The value of single validations is unclear, since there is substantial performance heterogeneity and good (or poor) performance on a single validation does not appear to reliably forecast performance on subsequent validations. No CPM showed consistently good discrimination across multiple validation databases. For example, the ten most validated CPMs have each been validated more than 20 times; all show substantial variation in discrimination across these validation studies, from virtually useless (i.e., c-statistic = ∼ 0.5) to very good (c-statistic = ∼0.8 or higher). This demonstrates the difficulty of defining the quality of a model generically, since performance greatly depends on characteristics of the database on which a model is tested. These findings underscore recent calls for a fundamental paradigm shift in how models are assessed for validity and utility^17^ and calls for more robust stewardship of algorithms for health care^18^.

The majority of cardiovascular CPMs in our Registry have never been externally validated. This finding mirrors an observation made in previous assessment of primary prevention models^8^ and broadly suggests that cardiovascular clinicians should be skeptical about the accuracy of individual risk estimates. In our Registry, model level predictors associated with subsequent external validation include the disease being studied and also larger sample size, higher outcome rates, and whether discrimination or calibration were reported in the original presentation. Older CPMs were generally more likely to be externally validated—an observation that may relate to insufficient time to allow for validation of more recently published CPMs. Given the extreme redundancy of CPMs and the relative scarcity of external validations, it seems reasonable to prioritize the study of existing cardiovascular CPMs (as opposed to developing new ones), and how these might be optimized for clinical use.

In our review, it was common to observe substantial decrements in discrimination during validations. This finding is consistent with prior reports that have shown CPM validation discriminatory ability that is highly variable and often worse than anticipated (when compared to performance on the derivation database).^6,8^ There are several potential reasons why model performance might decrease, including model invalidity (e.g., due to over-fitting on the derivation population) and a change in case mix.^5^ Model invalidity might be expected to be more pronounced when models are evaluated in populations that are dissimilar to the derivation population. We found that models had a substantially larger decrease in model performance when tested on distantly-related populations compared to either related or closely-related populations. However, judging the relatedness of the populations is laborious and requires substantial clinical expertise. Differences that may appear subtle can be very influential.

For example, a CPM developed on patients in the emergency room might not be expected to have similar discriminatory performance if the validation cohort includes only patients admitted to the hospital since—as in the case of many acute cardiac syndromes—care^19^ and outcome predictors^20^ are different very early in the disease course. So too changes in treatments received (e.g., different ACS revascularization approaches,^21^ stent types^22^, or outcome definitions^23,24^) likely impact model validation performance. If the model was derived on patients receiving lytic therapy and validated using data from a more contemporary percutaneous coronary intervention (PCI) trial, it should not be surprising that model performance appears worse than expected. Other study-level characteristics we examined apart from relatedness, did not appear to greatly influence model performance.

One of the most striking observations of this work is that isolated validations appear insufficient to understand the performance of CPMs when tested in new populations. There was often an extreme range in performance for CPMs evaluated in multiple databases—an observation that calls into question the generalizability of any one validation result. These data challenge the current approach in which a model might be evaluated on a single external population and then declared to be a ‘validated’ prediction model that is ready for use. Even when a model performs well using statistical criteria, it is unclear whether such a model improves decision making when used on a closely related population.

Further, good statistical performance on one external database does not guarantee good statistical performance in another setting—such as where a CPM is eventually used to support care. There is no evidence from our analysis that so called “validated” CPMs that have been integrated into clinical practice guidelines^25,26^ should be accepted as trustworthy unless CPM performance is specifically known to be excellent on populations like those being treated. While having a single CPM that is accepted by the clinical community and promoted in guidelines is appealing as a means of standardizing practice across a range of different settings, the degree of variation seen in our review suggests that this paradigm may result in substantial variation of performance across different settings, and poor performance in some settings. Testing CPMs for improved decision making and better clinical outcomes (e.g. in a cluster randomized trial^27^) is rarely performed prior to dissemination into practice. Novel paradigms, emphasizing increasing the accuracy of model performance on local populations, through continual recalibration and updating, are an appealing approach that deserves further consideration.

Our review has several limitations. First, the review was limited by the information collected and presented in the original articles. We relied on changes in discrimination largely because CPM calibration is woefully under-assessed. Only 62% of models in the CPM Registry have had calibration formally assessed in an external population; even among the models that were validated only 48% report any calibration. Finally, even when calibration is reported, it is usually reported in a form that is not clinically interpretable (e.g., as a Hosmer-Lemeshow statistic^4,13^) or graphically [easy to summarize according to calibration slope (ideal: 1) and systematic under or overestimation (intercept ideally 0)]. Some less frequently used metrics, such as the integrated calibration index^28^, may help compare performance across multiple validations. Decrements in calibration may be as serious as, or even more serious than, decrements in discrimination, since miscalibrated models yield misinformation which may cause harmful decision making.^9^ Ideally, we would be able to evaluate the ‘net benefit’ of model use, which integrates discrimination, calibration and relative utility to compare the value of prediction-based decision making compared to best “one-sized-fits-all” strategies.^4,29^ Such evaluations would have required individual patient data, since these approaches are so rarely used in the published literature. Similarly, we could not assess how much of the decrement in discrimination was due to differences in case mix, rather than invalidity, which would have also required evaluation of patient level data^30^. Finally, our systematic review does not include more recent validations after 2017, due to the enormous scope of this literature, the lack of efficient search strategies and the laborious nature of comprehensive data extraction and evaluation of relatedness. We do not anticipate the more recent literature would substantially change our findings. Maintenance and continual updating data of this registry will registry will require a semi-automated approach heavily reliant on natural language processing.^31^

## Conclusion

Many published cardiovascular CPMs have never been externally validated and for those that have, it is common to see significant performance heterogeneity and marked decreases in the discriminatory performance compared to the model development phase. Calibration has been widely under-assessed and single validations do not sufficiently capture CPM performance. Granular information about population relatedness is associated with CPM performance in external validations and when CPMs are tested on distantly related populations, model performance is often substantially worse than expected. This review raises substantial concerns about the current approach to ‘validating’ cardiovascular CPMs and underscores the need for a radical rethinking for how performance heterogeneity is explored and quantified (e.g. through multiple validations across various practice settings) and how models are evaluated for clinical use.

## Data Availability

The authors confirm that the data supporting the findings of this study are available within the article.

## Acknowledgements

Research reported in this work was funded through a Patient-Centered Outcomes Research Institute (PCORI) Award (ME-1606-35555). The views in this work are solely the responsibility of the authors and do not necessarily represent the views of the Patient-Centered Outcomes Research Institute (PCORI), its Board of Governors or Methodology Committee.

The authors wish to acknowledge the contributions of Mr. Vandan Patel for his work on the relatedness effort.

BW is supported by K23AG055667 from NIH-NIA and R03AG056447 from NIH-NIA

