## Supplemental Figure 2 for "External Validations of Cardiovascular Clinical Prediction Models: A Large-scale Review of the Literature"

### Supplement

**Supplemental Figure 2. Waterfall Plots by Condition**

A	B	C	D	E

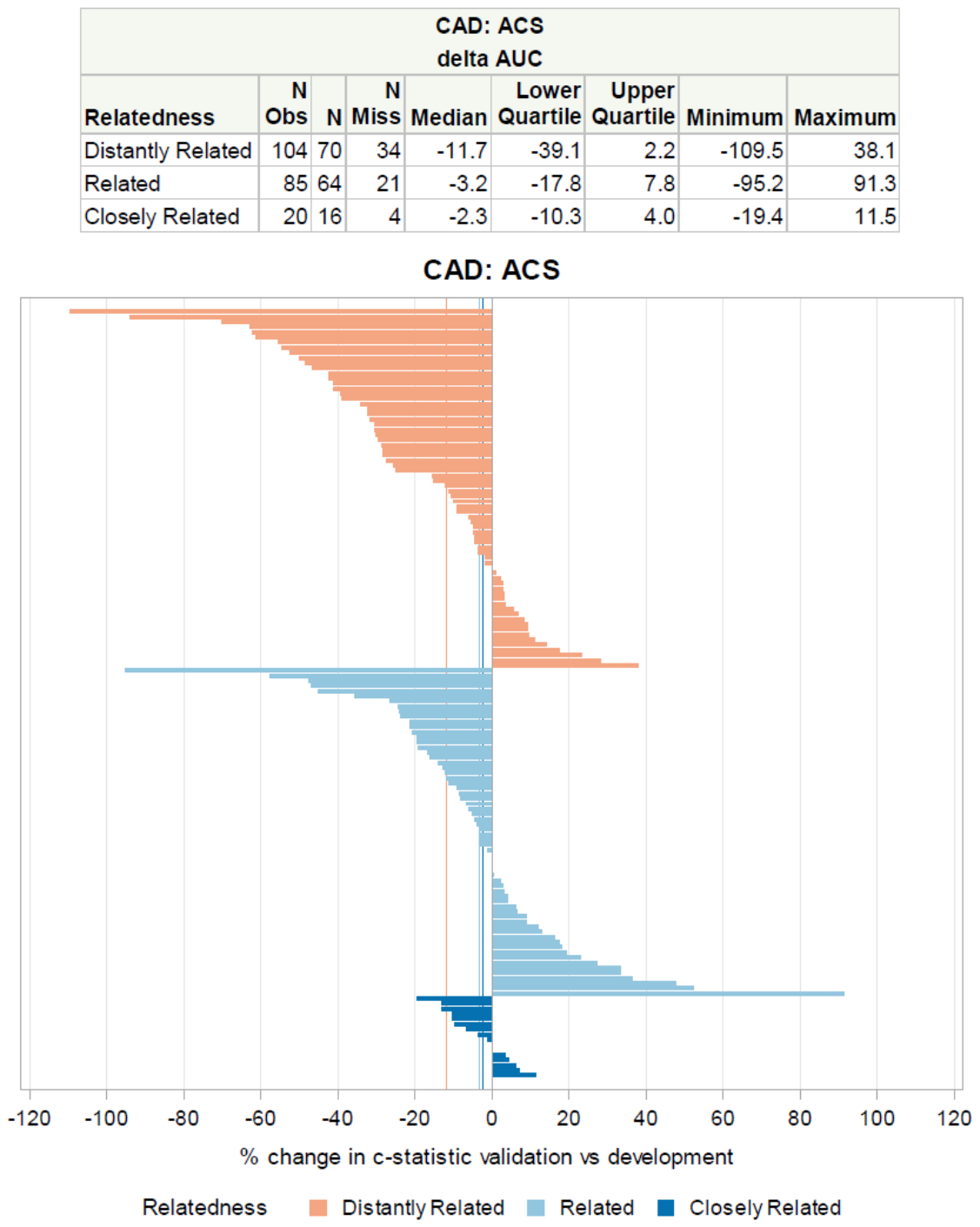

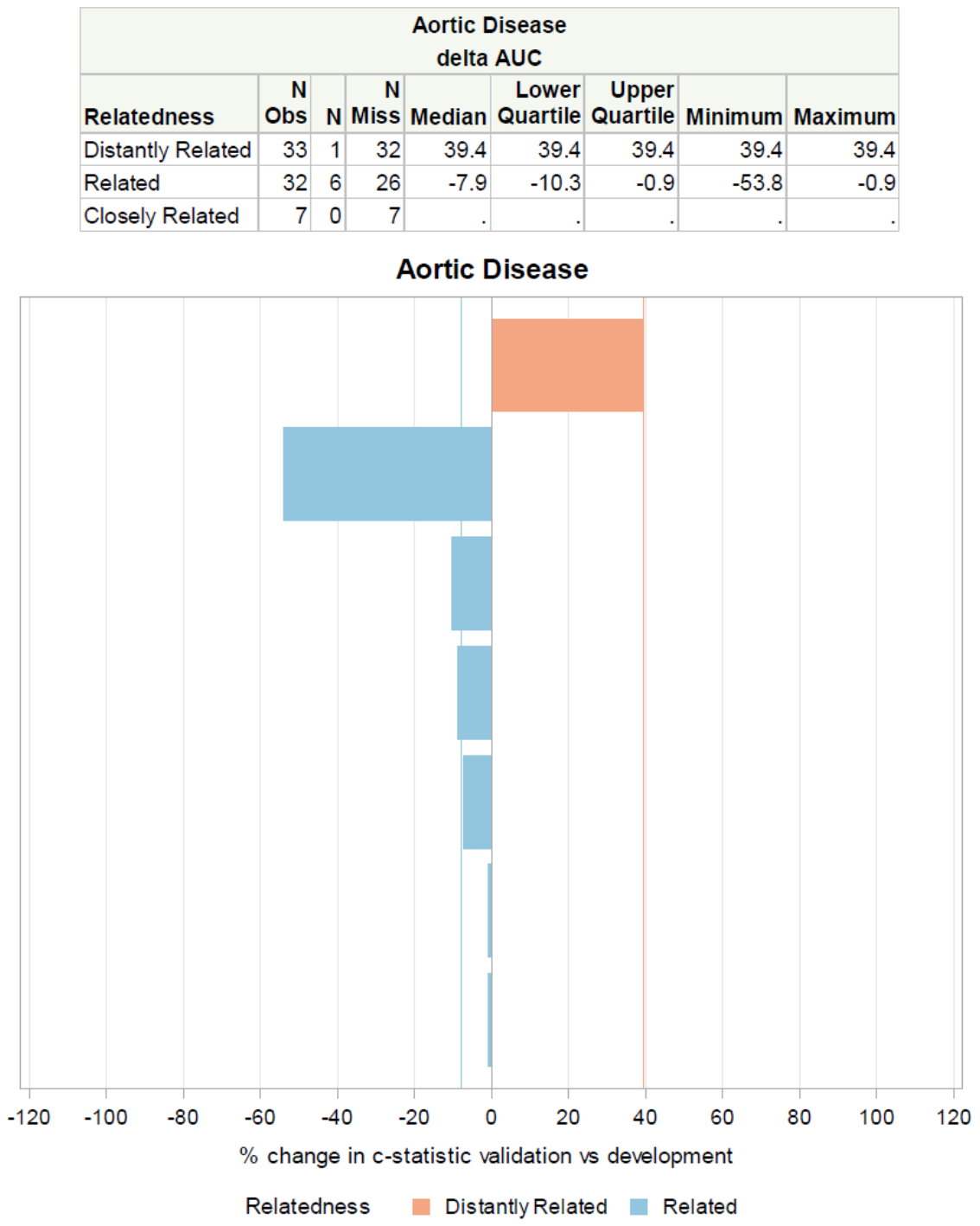

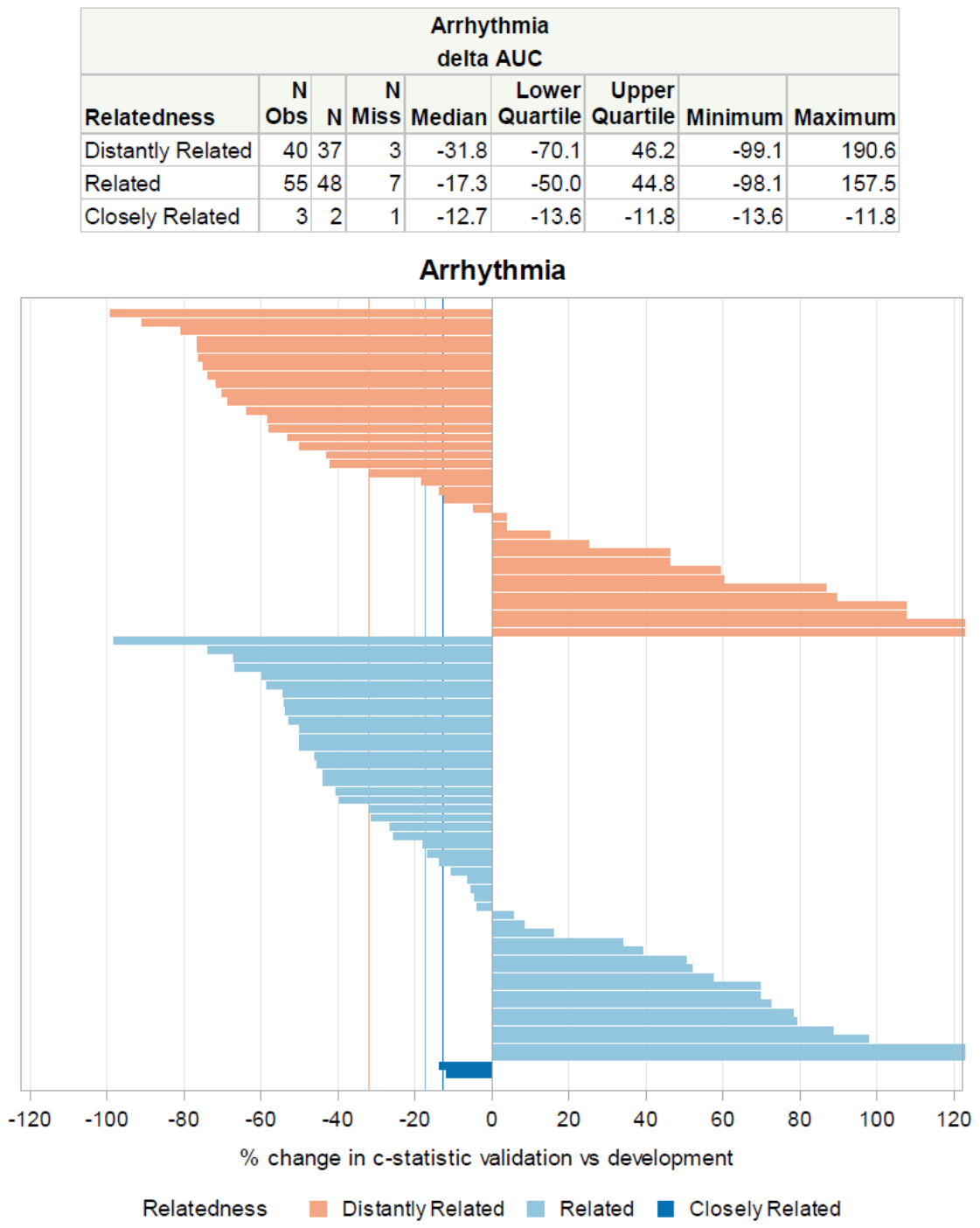

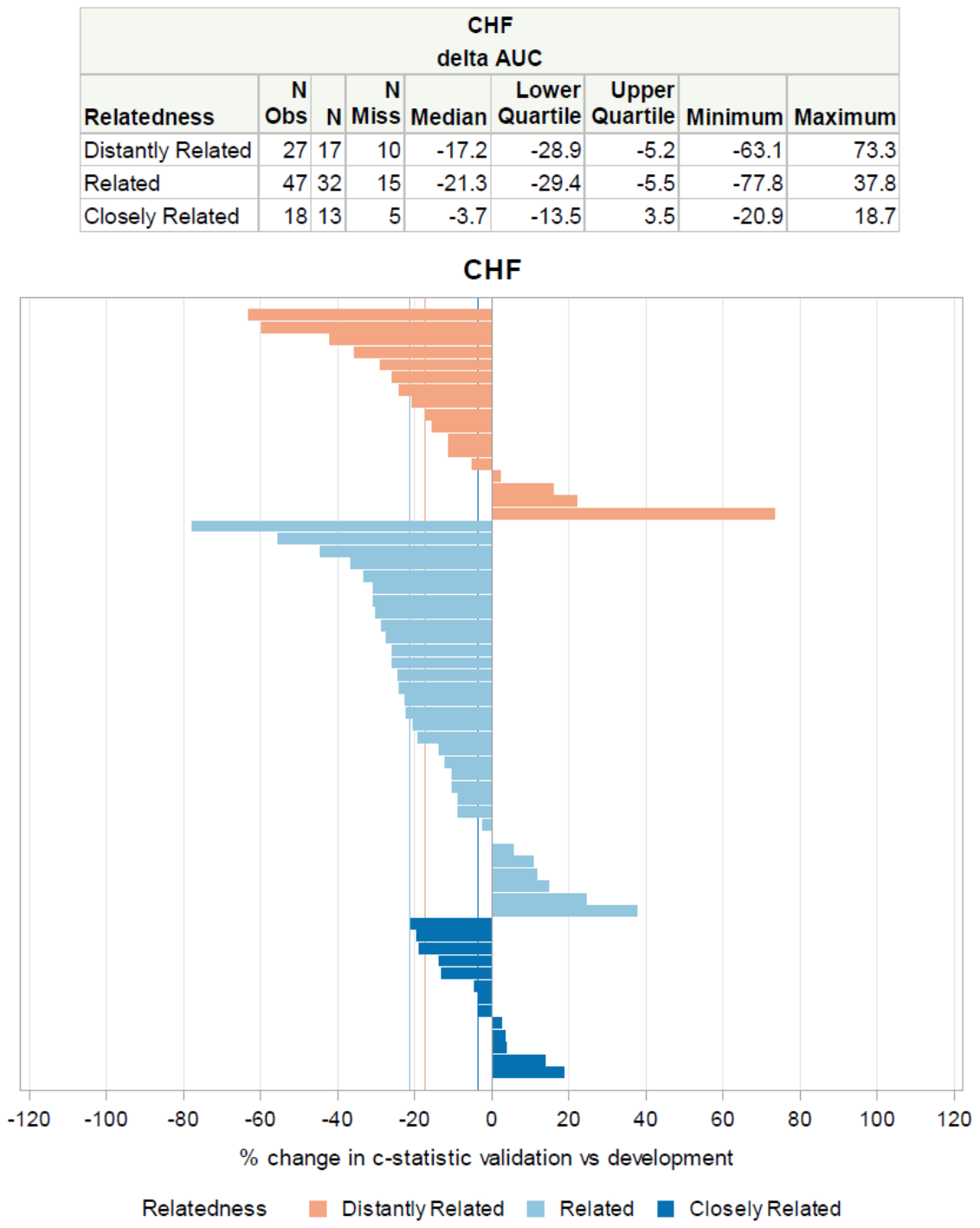

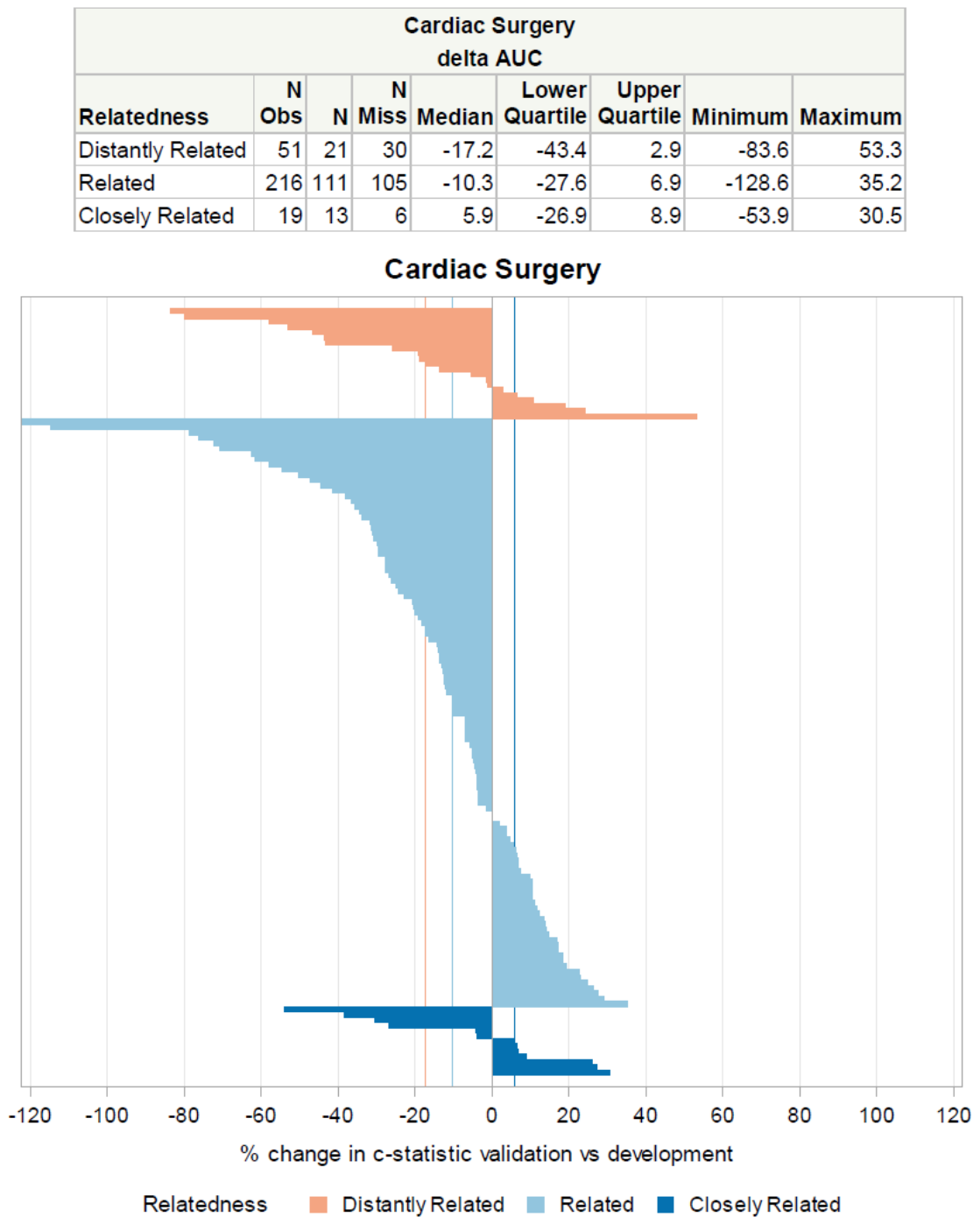

F	G	H	I	J

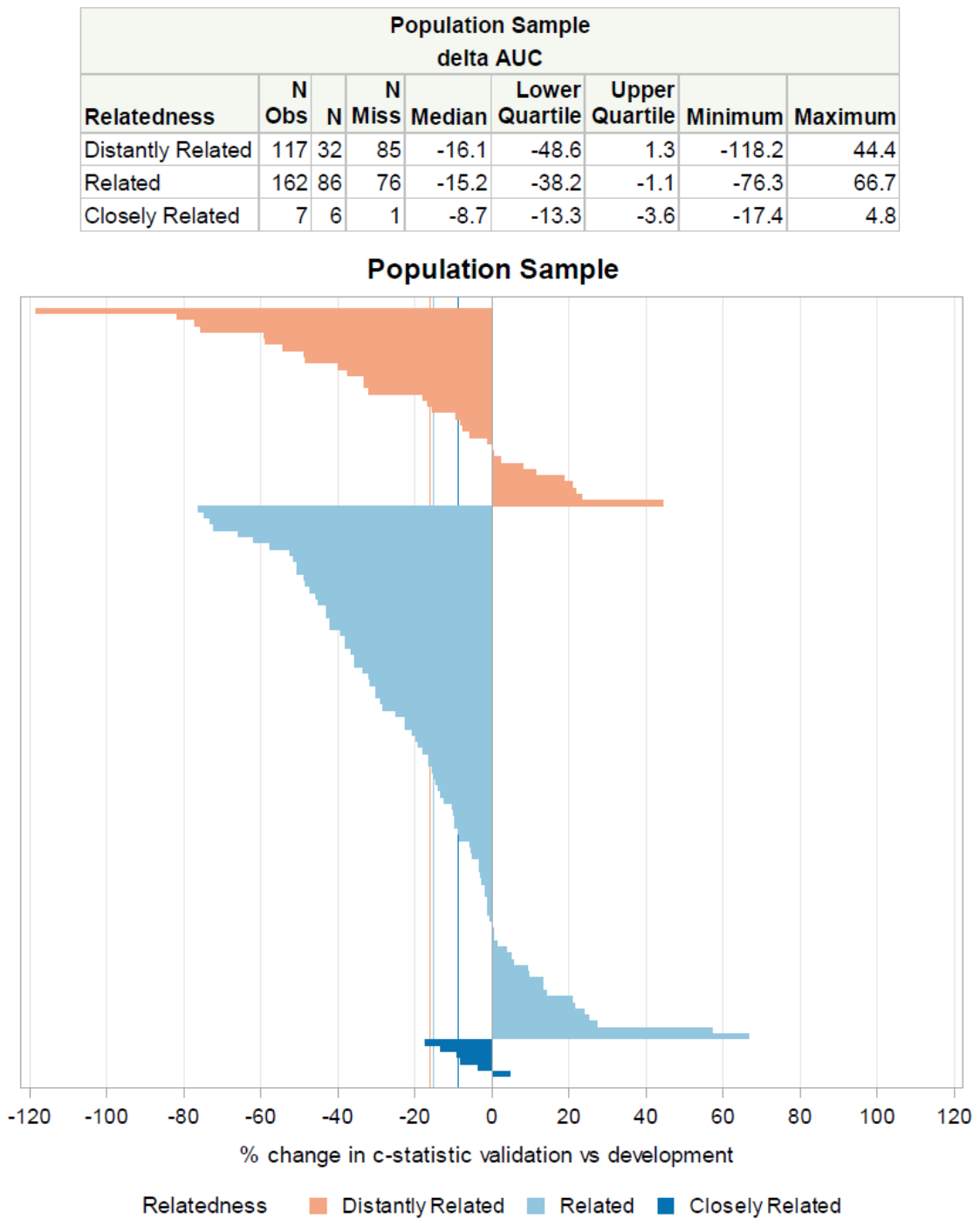

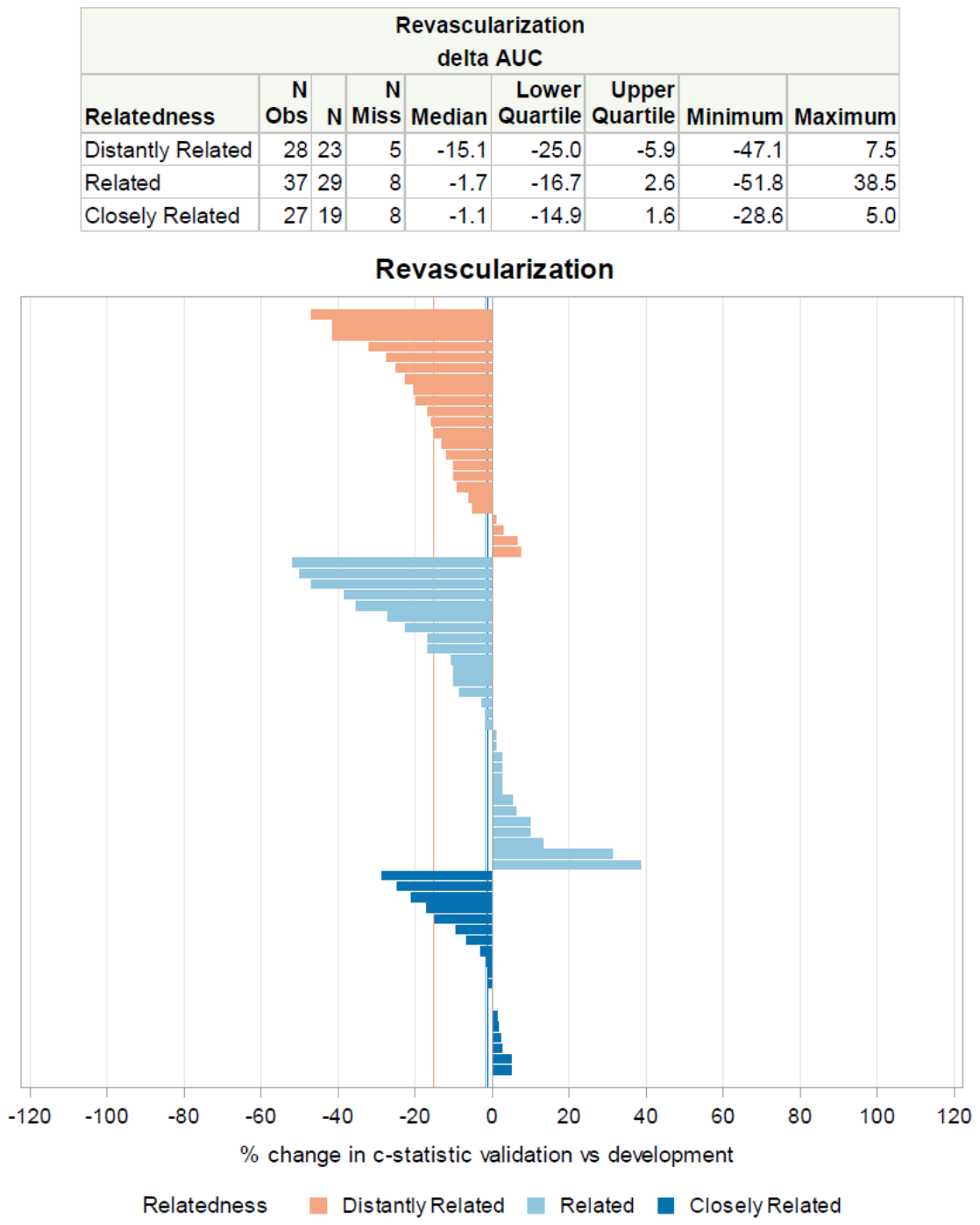

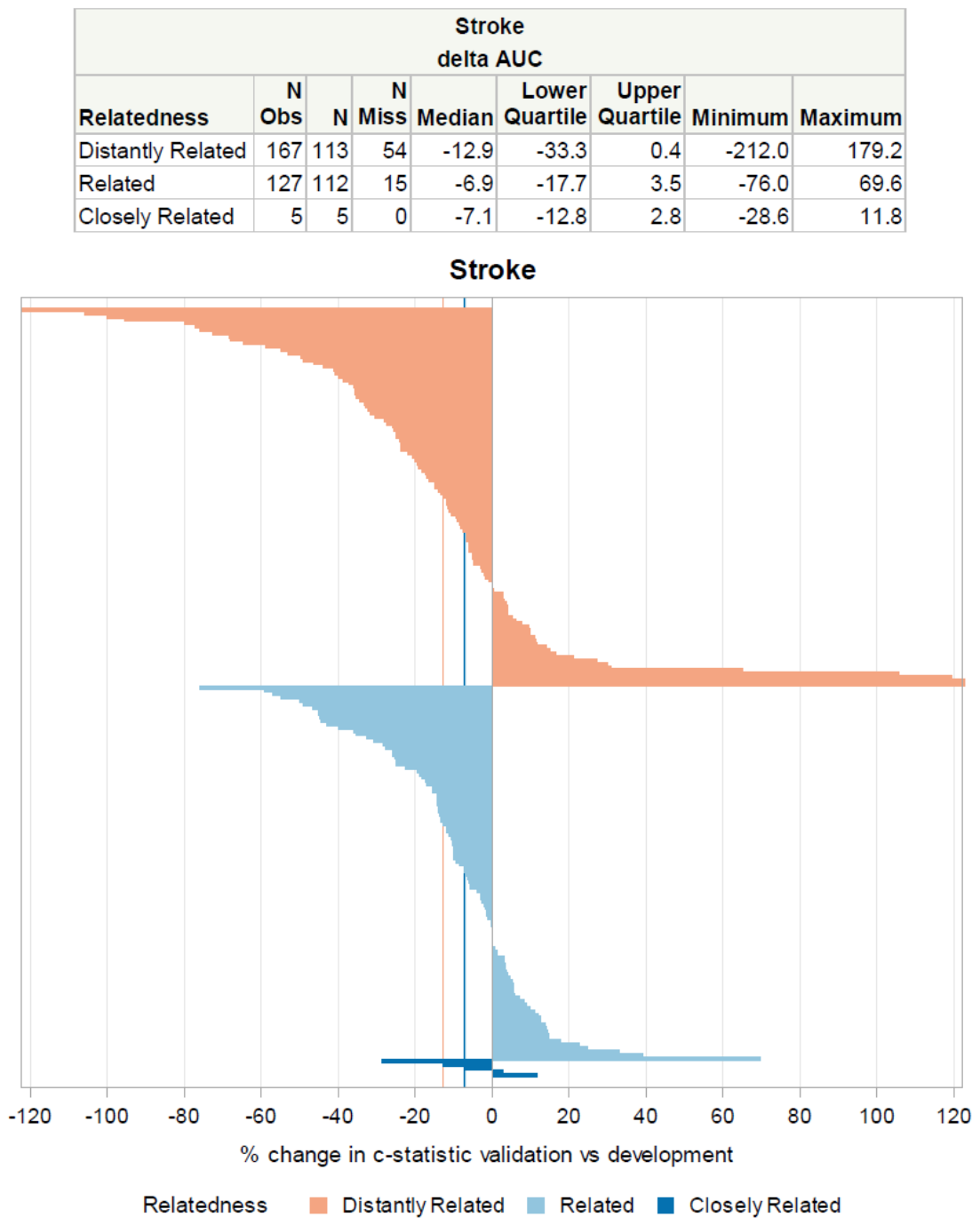

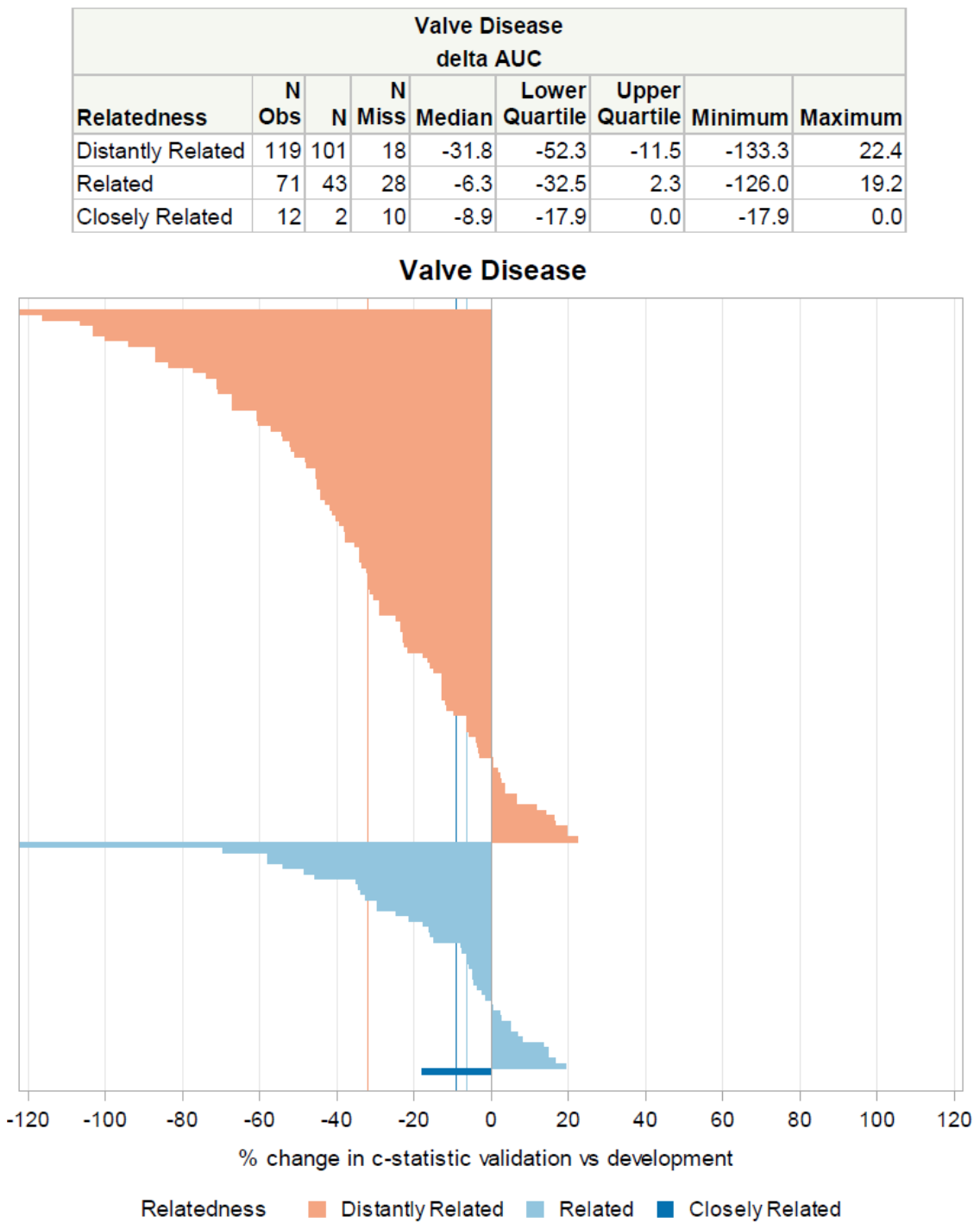

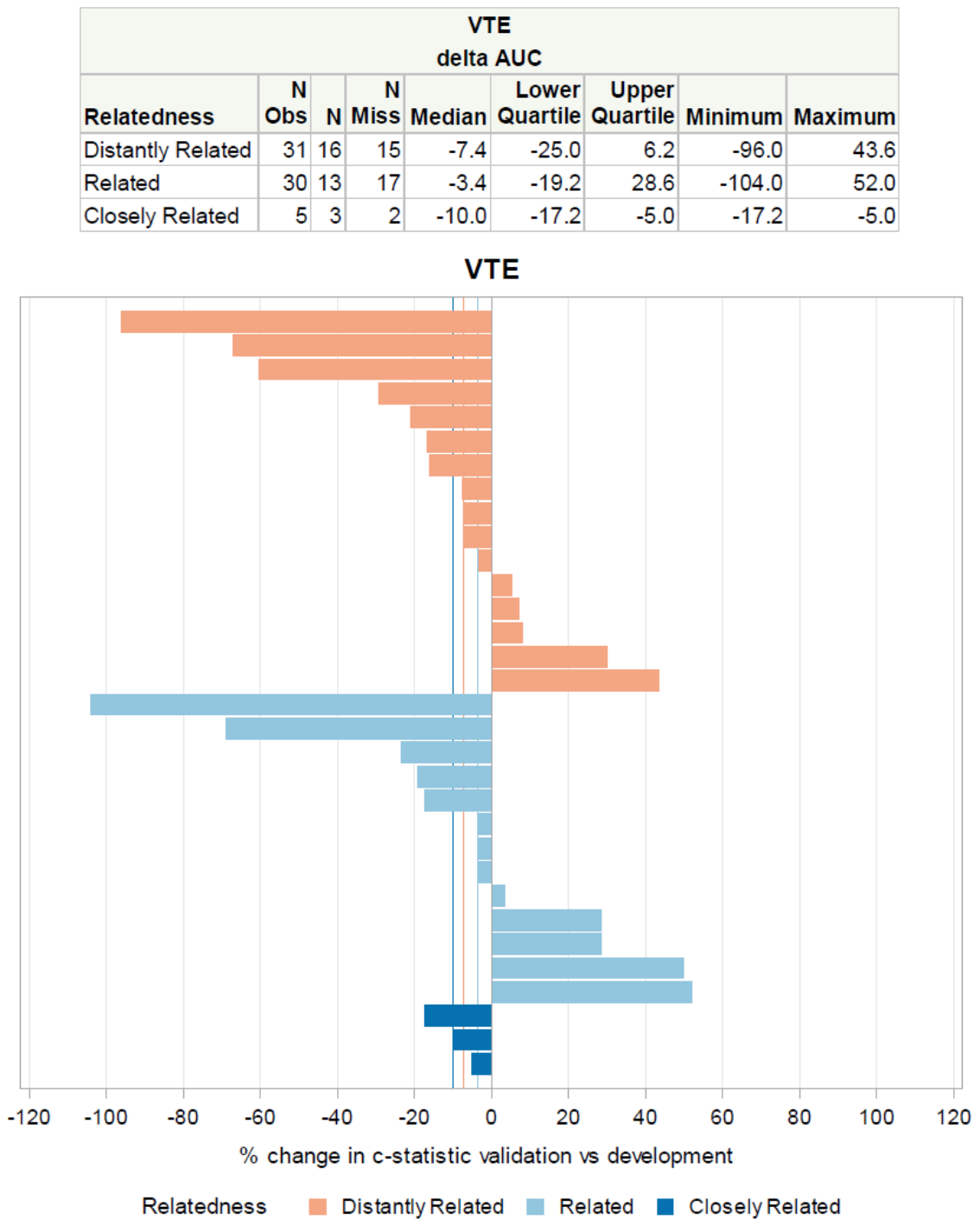

Waterfall plots depicting the percent change in the c-statistic in closely related validations (in dark blue), related validations (in light blue) and unrelated validations (in orange) for the top 10 most validated index conditions: A) ACS, B) Aortic Disease, C) Arrhythmia, D) Chronic Heart Failure, E) Cardiac Surgery, F) Population Sample, G) Revascularization, H) Stroke, I) Valve Disease, and J) Venous thromboembolism. Vertical lines show that the median decrement in discrimination was generally more pronounced in the distantly related models than the related or closely related models for 7 out of the 10 conditions shown above. Conditions for which there was not a pronounced decrement in performance had less than 100 validations.
