## Supplemental Table 1 for "External Validations of Cardiovascular Clinical Prediction Models: A Large-scale Review of the Literature"

### Supplement

**Supplemental Table 1. Relatedness Rubrics and Guidelines for the top 10 most validated index Conditions.**

| **Acute Coronary Syndrome (ACS)** | |
| --- | --- |
| **Domain** | **Guide** |
| **Split sample Check** | Split sample = Closely Related |
| **Index Condition Match** | No = Mismatch 🡪 Remove from sample |
| ***Recruitment Indication*** | |
| Inclusion Criteria | Partial match if no major clinical difference = Related |
| Exclusion Criteria |  |
| Recruitment Setting | If only major difference, ED vs. inpatient = Related |
| ACS Type | Partial match = Related |
| ***Intervention*** | |
| 1. CABG (%) | Partial match (+/- 30%) = Related |
| 2. PCI (%) |  |
| 3. Thrombolysis (%) |  |
| ***Study Characteristics*** | |
| Enrollment Period | Partial match (<20 difference) = Related |
| Follow-up time | If only difference,  1) Cox model = Related  2) Derivation time short enough to be logistic regression vs. validation is longer = Distantly Related |
| Outcome | No match = not validation 🡪 Remove from sample |
| ED indicated emergency department; CABG, coronary artery bypass graft; PCI, percutaneous coronary intervention. | |

| **Aortic Disease** | |
| --- | --- |
| **Domain** | **Guide** |
| **Split sample Check** | Split sample = Closely Related |
| **Index Condition Match** | No = Mismatch 🡪 Remove from sample |
| ***Recruitment Indication*** | |
| Inclusion Criteria | Partial match if no major clinical difference = Related |
| Exclusion Criteria |  |
| Recruitment Setting | If only major difference, ED vs. inpatient = Related |
| Aneurysm Site | Partial match = Related |
| Status | Elective vs. Non-elective = Distantly Related |
| ***Intervention*** | |
| Intervention | Partial match = Related |
| ***Study Characteristics*** | |
| Enrollment Period | Partial match (<20 difference) = Related |
| Follow-up time | If only difference,  1) Cox model = Related  2) Derivation time short enough to be logistic regression vs. validation is longer = Distantly Related |
| Outcome | No match = not validation 🡪 Remove from sample |
| ED indicated emergency department. | |

| **Arrhythmia** | |
| --- | --- |
| **Domain** | **Guide** |
| **Split sample Check** | Split sample = Closely Related |
| **Index Condition Match** | No = Mismatch 🡪 Remove from sample |
| ***Recruitment Indication*** | |
| Inclusion Criteria | Partial match if no major clinical difference = Related |
| Exclusion Criteria |  |
| Recruitment Setting | If only major difference, ED vs. inpatient = Related |
| Type of Arrhythmia | Partial match = Related |
| ***Intervention*** | |
| Intervention | Partial match = Related |
| ***Study Characteristics*** | |
| Enrollment Period | Partial match (<20 difference) = Related |
| Follow-up time | If only difference,  1) Cox model = Related  2) Derivation time short enough to be logistic regression vs. validation is longer = Distantly Related |
| Outcome | No match = not validation 🡪 Remove from sample |
| ED indicated emergency department. | |

| **Chronic Heart Failure (CHF)** | |
| --- | --- |
| **Domain** | **Guide** |
| **Split sample Check** | Split sample = Closely Related |
| **Index Condition Match** | No = Mismatch 🡪 Remove from sample |
| ***Recruitment Indication*** | |
| Inclusion Criteria | Partial match if no major clinical difference = Related |
| Exclusion Criteria |  |
| Recruitment Setting | If only major difference, ED vs. inpatient = Related |
| ***LVEF Subtype*** | |
| Reduced LVEF (%) | Partial match (+/- 30%) = Related |
| Preserved LVEF (%) |  |
| ***Population Cohort Disease Status*** | |
| ADHF (%) | Partial match (+/- 30%) = Related |
| Mean NYHA Class |  |
| CHF: NYHA Class III/IV) |  |
| CHF: NYHA Class I/II) |  |
| ***Medication*** | |
| 1. Beta Blockers (%) | Partial match (+/- 30%) = Related |
| 2. ACE inhibitors (%) |  |
| 3. ARB (%) |  |
| 4. ICD (%) |  |
| ***Study Characteristics*** | |
| Enrollment Period | Partial match (<20 difference) = Related |
| Follow-up time | If only difference,  1) Cox model = Related  2) Derivation time short enough to be logistic regression vs. validation is longer = Distantly Related |
| Outcome | No match = not validation 🡪 Removed from sample |
| ED indicated emergency department; LVEF, left ventricular ejection fraction; ADHF, acute decompensated heart failure; NYHA, New York Heart Association; ACE, angiotensin-converting enzyme; ARB, angiotensin receptor blocker; ICD, implantable cardioverter device. | |

| **Cardiac Surgery** | |
| --- | --- |
| **Domain** | **Guide** |
| **Split sample Check** | Split sample = Closely Related |
| **Index Condition Match** | No = Mismatch 🡪 Remove from sample |
| ***Recruitment Indication*** | |
| Inclusion Criteria | Partial match if no major clinical difference = Related |
| Exclusion Criteria |  |
| ***Surgical Procedure*** | |
| 1. Isolated CABG (%) | Partial match (+/- 30%) = Related |
| 2. Valve + CABG (%) |  |
| 3. CHD-related procedure (%) |  |
| 3. Other procedures (%) |  |
| ***Study Characteristics*** | |
| Enrollment Period | Partial match (<20 difference) = Related |
| Follow-up time | If only difference,  1) Cox model = Related  2) Derivation time short enough to be logistic regression vs. validation is longer = Distantly Related |
| Outcome | No match = not validation 🡪 Removed from sample |
| CABG indicates coronary artery bypass graft; CHD, congenital heart defect. | |

| **Population Sample** | |
| --- | --- |
| **Domain** | **Guide** |
| **Split sample Check** | Split sample = Closely Related |
| **Index Condition Match** | No = Distantly Related |
| ***Recruitment Indication*** | |
| Inclusion Criteria | Partial match if no major clinical difference = Related |
| Exclusion Criteria |  |
| Recruitment Setting | If only major difference, ED vs. inpatient = Related |
| ***Risk Factors*** | |
| 1. Diabetes (%) | Partial match (+/- 10%) = Related  BUT if Risk Factor part of inclusion criteria (i.e., 100% vs. other %) then >50% different = Distantly Related |
| 2. Hypertension (%) |  |
| 3. Hyperlipidemia (%) |  |
| ***Study Characteristics*** | |
| Enrollment Period | Partial match (<20 difference) = Related |
| Follow-up time | If only difference,  1) Cox model = Related  2) Derivation time short enough to be logistic regression vs. validation is longer = Distantly Related |
| Outcome | No match = not validation 🡪 Removed from sample |
| ED indicated emergency department. | |

| **Revascularization** | |
| --- | --- |
| **Domain** | **Guide** |
| **Split sample Check** | Split sample = Closely Related |
| **Index Condition Match** | No = Mismatch 🡪 Remove from sample |
| ***Recruitment Indication*** | |
| Inclusion Criteria | Partial match if no major clinical difference = Related |
| Exclusion Criteria |  |
| Recruitment Setting | If only major difference, ED vs. inpatient = Related |
| ***Study Characteristics*** | |
| Enrollment Period | Partial match (<20 difference) = Related |
| Follow-up time | If only difference,  1) Cox model = Related  2) Derivation time short enough to be logistic regression vs. validation is longer = Distantly Related |
| Outcome | No match = not validation 🡪 Removed from sample |
| ED indicated emergency department. | |

| **Stroke** | |
| --- | --- |
| **Domain** | **Guide** |
| **Split sample Check** | Split sample = Closely Related |
| ***Stroke*** | |
| Stroke Type | Partial match = Distantly Related |
| Treatment Type | Partial match = Distantly Related |
| Time since stroke onset | Partial match = Related |
| Timing of model use | Partial match = Related |
| ***Study Characteristics*** | |
| Enrollment Period | Partial match (<20 difference) = Related |
| Follow-up time | No match (Short time frame: >3 months; Long time frame: >9 months) = Distantly Related |
| Outcome | No match = not validation 🡪 Removed from sample |

| **Venous Thromboembolism (VTE)** | |
| --- | --- |
| **Domain** | **Guide** |
| **Split sample Check** | Split sample = Closely Related |
| **Index Condition Match** | No = Mismatch 🡪 Remove from sample |
| ***Recruitment Indication*** | |
| Inclusion Criteria | Partial match if no major clinical difference = Related |
| Exclusion Criteria |  |
| Recruitment Setting | If only major difference, ED vs. inpatient = Related |
| ***Risk Factors*** | |
| 1. Cancer (%) | Partial match (+/- 10%) = Related  BUT if Risk Factor part of inclusion criteria (i.e., 100% vs. other %) then >50% different = Distantly Related |
| 2. Recent Surgery and/or Immobilization (%) |  |
| 3. Obstetrics (%) |  |
| ***Study Characteristics*** | |
| Enrollment Period | Partial match (<20 difference) = Related |
| Follow-up time | If only difference,  1) Cox model = Related  2) Derivation time short enough to be logistic regression vs. validation is longer = Distantly Related |
| Outcome | No match = not validation 🡪 Removed from sample |
| ED indicated emergency department. | |

| **Valve Disease** | |
| --- | --- |
| **Domain** | **Guide** |
| **Split sample Check** | Split sample = Closely Related |
| **Index Condition Match** | No = Mismatch 🡪 Remove from sample |
| ***Risk Factors*** | |
| 1. Intervention | Partial match (+/- 30%) = Related |
| 2. Revascularization/Stent (% Valve) | >10% difference = Distantly Related |
| ***Study Characteristics*** | |
| Continent | Partial match if no major clinical difference = Related |
| Enrollment Period | Non-overlapping treatment years = Distantly Related |
| Outcome | No match = not validation 🡪 Removed from sample |
