## Supplemental Table 2 for "External Validations of Cardiovascular Clinical Prediction Models: A Large-scale Review of the Literature"

### Supplement

**Supplemental Table 2. Predictors of External Validation.**

| **LR Statistics For global p-value** | | | | **Analysis Of Maximum Likelihood Parameter Estimates** | | | | | | | | |
| --- | --- | --- | --- | --- | --- | --- | --- | --- | --- | --- | --- | --- |
| **Source** | **DF** | **Chi-Square** | **Pr > ChiSq** | **Parameter** | | **Estimate** | **SE** | **Wald ChiSq** | **Pr > ChiSq** | **Odds ratio** | **Lower 95%CI** | **Upper 95%CI** |
| Index condition | 18 | 90.29 | <.0001 | *Intercept* | | -1.6352 | 0.384 | 18.13 | <.0001 |  |  |  |
|  |  |  |  | **Index condition** | | | | | | | | |
| Internally validated | 1 | 31.57 | <.0001 | Population Sample | | *Reference* | - | - | - | - | - | - |
|  |  |  |  | Aortic Disease | | 0.9733 | 0.3727 | 6.82 | 0.009 | 2.65 | 1.27 | 5.49 |
| Publication year | 3 | 10.88 | 0.0124 | Arrhythmia | | 2.2642 | 0.5739 | 15.56 | <.0001 | 9.62 | 3.12 | 29.64 |
|  |  |  |  | CAD | | 2.1319 | 0.746 | 8.17 | 0.0043 | 8.43 | 1.95 | 36.38 |
| Continent | 5 | 14.55 | 0.0125 | CAD: ACS | | 0.2894 | 0.2888 | 1 | 0.3163 | 1.34 | 0.76 | 2.35 |
|  |  |  |  | CAD: Cardiac Surgery | | 1.0372 | 0.7259 | 2.04 | 0.1531 | 2.82 | 0.68 | 11.70 |
| Study Design | 2 | 1.55 | 0.4617 | CAD: Stable | | -1.1416 | 0.4626 | 6.09 | 0.0136 | 0.32 | 0.13 | 0.79 |
|  |  |  |  | CHF | | 0.0883 | 0.2846 | 0.1 | 0.7563 | 1.09 | 0.63 | 1.91 |
| Sample size | 4 | 15.97 | 0.0031 | Cardiac Arrest | | -0.4357 | 0.5379 | 0.66 | 0.4179 | 0.65 | 0.23 | 1.86 |
|  |  |  |  | Cardiac Surgery | | 0.6747 | 0.3137 | 4.62 | 0.0315 | 1.96 | 1.06 | 3.63 |
| Number of events | 4 | 5.71 | 0.2216 | Carotid Disease | | -0.3706 | 0.6074 | 0.37 | 0.5418 | 0.69 | 0.21 | 2.27 |
|  |  |  |  | Chest Pain | | 0.5042 | 0.3988 | 1.6 | 0.2061 | 1.66 | 0.76 | 3.62 |
| Number of predictors | 1 | 6.26 | 0.0123 | Other | | 0.7235 | 0.37 | 3.82 | 0.0505 | 2.06 | 1.00 | 4.26 |
|  |  |  |  | PVD | | -0.7762 | 0.8222 | 0.89 | 0.3451 | 0.46 | 0.09 | 2.31 |
| Prediction horizon | 4 | 5.26 | 0.262 | Post-Operative | | -0.514 | 0.7244 | 0.5 | 0.478 | 0.60 | 0.14 | 2.47 |
|  |  |  |  | Revascularization | | 0.1152 | 0.2983 | 0.15 | 0.6995 | 1.12 | 0.63 | 2.01 |
| Regression method | 3 | 1.46 | 0.6926 | Stroke | | 1.0989 | 0.2766 | 15.79 | <.0001 | 3.00 | 1.75 | 5.16 |
|  |  |  |  | VTE | | 1.1949 | 0.3734 | 10.24 | 0.0014 | 3.30 | 1.59 | 6.87 |
| Discrimination reported | 1 | 5.11 | 0.0238 | Valve Disease | | 1.1388 | 0.3663 | 9.67 | 0.0019 | 3.12 | 1.52 | 6.40 |
|  |  |  |  | **CPM Development Parameter** | | | | | | | | |
| Calibration reported | 1 | 5.06 | 0.0244 | Internally validated | |  |  |  |  |  |  |  |
|  |  |  |  |  | Yes vs no | -0.8129 | 0.1476 | 30.33 | <.0001 | 0.44 | 0.33 | 0.59 |
|  |  |  |  | Publication year | |  |  |  |  |  |  |  |
|  |  |  |  |  | Q1 group (before 2004) | *Reference* | - | - | - | - | - | - |
|  |  |  |  |  | Q2 group (2004-2009) | -0.1836 | 0.1729 | 1.13 | 0.2883 | 0.83 | 0.59 | 1.17 |
|  |  |  |  |  | Q3 group (2009-2012) | -0.5817 | 0.1951 | 8.89 | 0.0029 | 0.56 | 0.38 | 0.82 |
|  |  |  |  |  | Q4 group (after 2012) | -0.5717 | 0.2094 | 7.45 | 0.0063 | 0.56 | 0.37 | 0.85 |
|  |  |  |  | Continent | |  |  |  |  |  |  |  |
|  |  |  |  |  | Europe | *Reference* | - | - | - | - | - | - |
|  |  |  |  |  | Asia | -0.5444 | 0.487 | 1.25 | 0.2636 | 0.58 | 0.22 | 1.51 |
|  |  |  |  |  | Australia | 1.0921 | 0.4728 | 5.34 | 0.0209 | 2.98 | 1.18 | 7.53 |
|  |  |  |  |  | International | 0.1069 | 0.4041 | 0.07 | 0.7913 | 1.11 | 0.50 | 2.46 |
|  |  |  |  |  | North America | 0.5681 | 0.2367 | 5.76 | 0.0164 | 1.76 | 1.11 | 2.81 |
|  |  |  |  |  | Not reported | 0.3367 | 0.1523 | 4.89 | 0.0271 | 1.40 | 1.04 | 1.89 |
|  |  |  |  | Study design | |  |  |  |  |  |  |  |
|  |  |  |  |  | RCT | *Reference* | - | - | - | - | - | - |
|  |  |  |  |  | Cohort | -0.0916 | 0.2126 | 0.19 | 0.6664 | 0.91 | 0.60 | 1.38 |
|  |  |  |  |  | Other | -0.58 | 0.4755 | 1.49 | 0.2226 | 0.56 | 0.22 | 1.42 |
|  |  |  |  | Sample size | |  |  |  |  |  |  |  |
|  |  |  |  |  | Q1 group (<507) | *Reference* | - | - | - | - | - | - |
|  |  |  |  |  | Q2 group (507 - <1728) | 0.6562 | 0.1912 | 11.78 | 0.0006 | 1.93 | 1.33 | 2.80 |
|  |  |  |  |  | Q3 group (172 - <6213) | 0.6452 | 0.2135 | 9.13 | 0.0025 | 1.91 | 1.25 | 2.90 |
|  |  |  |  |  | Q4 group (≥6213) | 0.8093 | 0.2553 | 10.05 | 0.0015 | 2.25 | 1.36 | 3.71 |
|  |  |  |  |  | Not reported | 0.0571 | 0.582 | 0.01 | 0.9218 | 1.06 | 0.34 | 3.31 |
|  |  |  |  | Events | |  |  |  |  |  |  |  |
|  |  |  |  |  | Q1 group (<71) | *Reference* | - | - | - | - | - | - |
|  |  |  |  |  | Q2 group (71 - <165) | 0.167 | 0.1936 | 0.74 | 0.3884 | 1.18 | 0.81 | 1.73 |
|  |  |  |  |  | Q3 group (165 - <456) | 0.2195 | 0.2153 | 1.04 | 0.3079 | 1.25 | 0.82 | 1.90 |
|  |  |  |  |  | Q4 group (≥456) | 0.5361 | 0.2493 | 4.63 | 0.0315 | 1.71 | 1.05 | 2.79 |
|  |  |  |  |  | Not reported | 0.4074 | 0.2283 | 3.18 | 0.0744 | 1.50 | 0.96 | 2.35 |
|  |  |  |  | Predictors (continuous) | |  |  |  |  |  |  |  |
|  |  |  |  |  | Per additional predictor | 0.0156 | 0.0064 | 5.9 | 0.0152 | 1.02 | 1.00 | 1.03 |
|  |  |  |  | Prediction horizon | |  |  |  |  |  |  |  |
|  |  |  |  |  | Short (<30 days) | *Reference* | - | - | - | - | - | - |
|  |  |  |  |  | Intermediate (30-365) | 0.2453 | 0.3076 | 0.64 | 0.4253 | 1.28 | 0.70 | 2.34 |
|  |  |  |  |  | Long (>365) | 0.03 | 0.2442 | 0.02 | 0.9022 | 1.03 | 0.64 | 1.66 |
|  |  |  |  |  | N/A | -0.0661 | 0.3279 | 0.04 | 0.8402 | 0.94 | 0.49 | 1.78 |
|  |  |  |  |  | Not reported | -0.7907 | 0.391 | 4.09 | 0.0432 | 0.45 | 0.21 | 0.98 |
|  |  |  |  | Regression method | |  |  |  |  |  |  |  |
|  |  |  |  |  | Logistic | *Reference* | - | - | - | - | - | - |
|  |  |  |  |  | Time-to-event | 0.1013 | 0.1799 | 0.32 | 0.5735 | 1.11 | 0.78 | 1.57 |
|  |  |  |  |  | Other | -0.1103 | 0.307 | 0.13 | 0.7194 | 0.90 | 0.49 | 1.63 |
|  |  |  |  |  | Not reported | 0.3699 | 0.3893 | 0.9 | 0.342 | 1.45 | 0.67 | 3.10 |
|  |  |  |  | Discrimination reported | |  |  |  |  |  |  |  |
|  |  |  |  |  | Yes vs no | 0.3187 | 0.1414 | 5.08 | 0.0242 | 1.38 | 1.04 | 1.81 |
|  |  |  |  | Calibration reported | |  |  |  |  |  |  |  |
|  |  |  |  |  | Yes vs no | 0.3096 | 0.1379 | 5.04 | 0.0247 | 1.36 | 1.04 | 1.79 |
| Among the n=1382 CPMs 575 were externally validated. The results shown are estimated from a multivariable logistic regression model predicting the probability of a CPM being externally validated.  LR indicates logistic regression; DF, degrees of freedom; Pr, probability; SE, standard error; CI, confidence interval; CAD, coronary artery disease; ACS, acute coronary syndrome; CHF, chronic heart failure; PVD, peripheral vascular disease; VTE venous thromboembolism; CPM, clinical prediction model; vs, versus; RCT, randomized clinical trial; N/A, not applicable. | | | | | | | | | | | | |
